# In-context learning for data-efficient classification of diabetic retinopathy with multimodal foundation models

**DOI:** 10.1101/2025.03.09.25323618

**Authors:** Murat S. Ayhan, Ariel Y. Ong, Eden Ruffell, Siegfried K. Wagner, David A. Merle, Pearse A. Keane

## Abstract

**Importance:** In-context learning, a prompt-based learning mechanism that enables multimodal foundation models to adapt to new tasks, can eliminate the need for retraining or large annotated datasets. We use diabetic retinopathy detection as an exemplar to probe in-context learning for ophthalmology.

**Objective:** To evaluate whether in-context learning using a multimodal foundation model (Google Gemini 1.5 Pro) can match the performance of a domain-specific model (RETFound) fine-tuned for diabetic retinopathy detection from color fundus photographs.

**Design/Setting/Participants:** This cross-sectional study compared two approaches for adapting foundation models to diabetic retinopathy detection using a public dataset of 516 color fundus photographs. The images were dichotomized into two groups based on the presence or absence of any signs of diabetic retinopathy. RETFound was fine-tuned for this binary classification task, while Gemini 1.5 Pro was assessed for it under zero-shot and few-shot prompting scenarios, with the latter incorporating random or k-nearest-neighbors-based sampling of a varying number of example images. For experiments, data were partitioned into training, validation, and test sets in a stratified manner, with the process repeated for 10-fold cross-validation.

**Main Outcome(s) and Measure(s):** Performance was assessed via accuracy, F1 score, and expected calibration error of predictive probabilities. Statistical significance was evaluated using Wilcoxon tests.

**Results:** The best in-context learning performance with Gemini 1.5 Pro yielded an average accuracy of 0.841 (95% CI: 0.803–0.879), F1 score of 0.876 (95% CI: 0.844–0.909), and calibration error of 0.129 (95% CI: 0.107–0.152). RETFound achieved an average accuracy of 0.849 (95% CI: 0.813–0.885), F1 score of 0.883 (95% CI: 0.852–0.915), and calibration error of 0.081 (95% CI: 0.066–0.097). While accuracy and F1 scores were comparable (p>0.3), RETFound’s calibration was superior (p=0.004).

**Conclusions and Relevance:** Gemini 1.5 Pro with in-context learning demonstrated performance comparable to RETFound for binary diabetic retinopathy detection, illustrating how future medical artificial intelligence systems may build upon such frontier models rather than being bespoke solutions.

**Key Points:** *Question:* Can in-context learning using a general-purpose foundation model (Gemini 1.5 Pro) achieve performance comparable to a domain-specific model (RETFound) for binary diabetic retinopathy detection from color fundus photographs?

*Findings:* In this cross-sectional study, Gemini 1.5 Pro demonstrated accuracy and F1 scores comparable to the fine-tuned RETFound model. While classification performance was similar, RETFound showed better calibration.

*Meaning:* In-context learning with general-purpose foundation models like Gemini 1.5 Pro offers a promising, accessible approach for diabetic retinopathy detection, potentially enabling broader clinical adoption of advanced AI tools without the need for retraining or large labeled datasets.

## Introduction

Foundation models trained on broad, multimodal data have demonstrated impressive performance in diverse scenarios (1–5) and hold promise as general-purpose solutions adaptable to various clinical tasks (6). However, their deployment in specialized domains like medical imaging often requires resource-intensive fine-tuning on domain-specific datasets, limiting accessibility and scalability in all settings, not just in resource-constrained ones (7).

Initially observed in natural language processing (8) and later in computer vision (9,10), in-context learning (ICL) offers an alternative to transfer learning that typically requires fine-tuning of a pretrained model to new tasks before making predictions. Unlike transfer learning via fine-tuning, ICL does not require model retraining or parameter updates and instead allows foundation models to be adapted to new tasks by conditioning them via task-specific prompts (11). These prompts typically include concise task descriptions along a few illustrative examples (12).

ICL is particularly relevant for medical use cases, as language-based prompting allows non-technical users to easily adjust a model’s behavior. By eliminating the need for extensive labeled datasets, computational resources, or coding expertise, ICL can expand access to cutting-edge AI, allowing clinicians and researchers to leverage powerful models developed outside healthcare without the burden of building custom solutions from scratch.

Recent reports of widespread adoption of the DeepSeek models (13) in Chinese hospitals exemplify this trend (14). Additionally, ICL can potentially enhance explainability by exploiting multimodal foundation models’ ability to generate natural language-based descriptions of clinically relevant features and decision rationales, aligning AI outputs with the interpretability needs of clinicians and stakeholders.

Recently, ICL has been shown to enable classification of pathology images while attaining performance comparable to or surpassing that of fine-tuned foundation models, despite using significantly fewer annotated examples (15). Given that ophthalmology is an imaging-driven specialty of medicine, we explore the potential of ICL in this field by applying it to a clinically relevant task: classification of diabetic retinopathy (DR) using color fundus photographs (CFPs). We demonstrate that ICL using a multimodal foundation model (Gemini 1.5 Pro) can achieve diagnostic performance comparable to RETFound, a state-of-the-art foundation model specialized for retinal imaging (1), while also providing fairly well-calibrated predictive uncertainty estimates simply via prompt engineering. In addition, we provide evidence that the Gemini model can offer a window into its decision mechanism through counterfactual reasoning, achieved solely through prompt engineering.

## Methods

### Dataset

We evaluated the performance of ICL against transfer learning using a well-known publicly available dataset, the Indian Diabetic Retinopathy Image Dataset (IDRiD) (16). 516 macula-centered images were acquired in mydriasis via a Kowa VX-10α digital fundus camera with a 50-degree field of view, and were taken from real-world examinations performed at an eye clinic in India (17). All images in the dataset were graded according to the International Clinical Diabetic Retinopathy Severity Scale (18) by two medical experts who provided adjudicated consensus grades (17). No demographic information, e.g., age, sex or ethnicity, was available in the public dataset.

**Table 1.** Summary of the IDRiD dataset.

| Labels | (N / %) |
| --- | --- |
| 0: No Apparent DR | 168 / 32.56 |
| 1: Mild Non-Proliferative DR | 25 / 4.85 |
| 2: Moderate Non-Proliferative DR | 168 / 32.56 |
| 3: Severe Non-Proliferative DR | 93 / 18.02 |
| 4: Proliferative DR | 62 / 12.02 |
|  | 516 / 100.00 |

### Diabetic Retinopathy Detection as a Binary Classification Task

To test ICL as a potential tool for DR detection, we defined our task as a binary classification problem by dichotomizing the severity labels into the following groups: {0} vs {1,2,3,4}. Thus, in the presence of any signs of DR, a classifier is expected to assign the positive class label 1 (DR present). Otherwise, it should assign 0, the negative class label indicating an absence of DR. For probabilistic classification, this can be achieved by estimating *p*(*y* = 1|*x*), where x is an image and the model, *f*(*x*), essentially outputs the probability of the image belonging to the positive class, i.e., y=1. Then, a simple thresholding scheme yields the most likely class label: If *p*(*y* = 1|*x*) ≥ 0. 5 then 1 else 0.

A well-calibrated classifier provides probability estimates that accurately reflect the true likelihood of its predictions being correct. This ensures that its outputs can be interpreted as confidence values, making it easier to assess the reliability of automated decisions. Such classifiers can be integrated into clinical workflows (19), aid in decision referrals, and highlight cases where clinical decision-making may be particularly challenging (20–22). Expected calibration error (ECE) summarizes the overall calibration quality of a classifier into a single metric by capturing the gap between its confidence and accuracy (23).

### Model Development

We induced classifiers to perform the binary DR detection task via two approaches: transfer learning and ICL (Fig. 1). Our code, including prompts tailored to the task, is also available at https://github.com/msayhan/ICL-Ophthalmology-Public.

**Figure 1:**
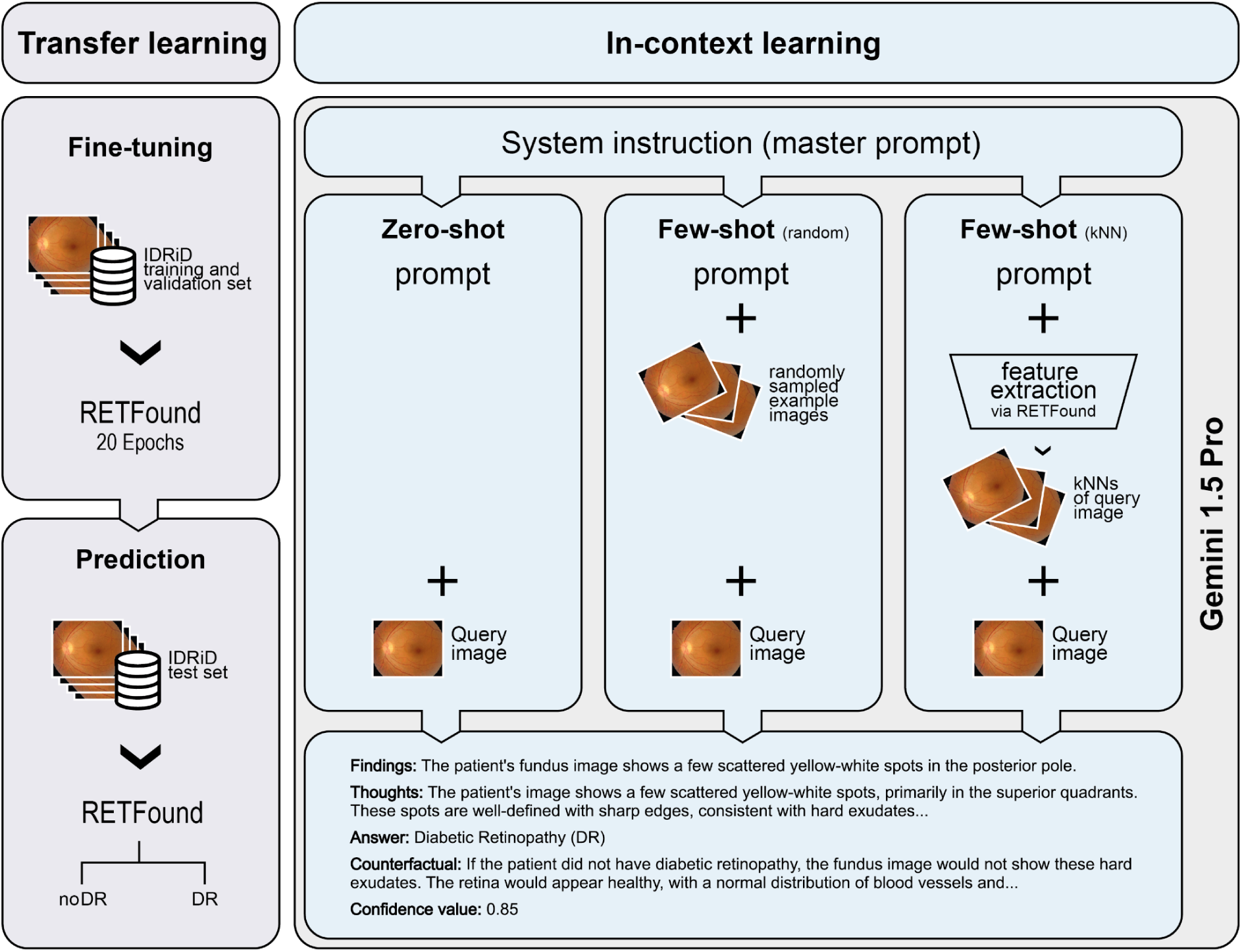
Study workflow. We performed binary DR classification using the IDRiD dataset. Transfer learning with RETFound served as a comparator (left panel). We used in-context learning with Gemini 1.5 Pro (right panel), employing three distinct prompting strategies: zero-shot prompting, few-shot prompting with randomly selected example images and few-shot prompting with k-nearest-neighbors (kNN)-based example selection. Across all in-context learning (ICL) scenarios, a system instruction (master prompt) was provided to define the model’s role. For details on dataset splitting and training procedures, refer to the methodological explanations provided in the Methods section.

#### Transfer learning

For transfer learning, we used RETFound (1), which was pretrained firstly on ImageNet (24) and then on 904,170 CFPs. We coupled its feature extraction encoder with a single linear layer for binary classification. In order to fine-tune and evaluate the model on IDRiD, we performed 10-fold stratified cross validation. Specifically, we allocated 10% of data for testing, while another 10% went for validation and the remaining 80% for training. We trained each model for 20 epochs using binary cross entropy loss, evaluating on the validation set after each epoch. We then selected the models with the smallest validation loss for testing.

During training, we used a weight decay parameter of 0.01, an initial learning rate of 0.001, which is linearly scaled with the minibatch size of 8 times batch accumulation steps of 2 divided by 256, and a layer decay of 0.75. Our optimizer was AdamW (25) coupled with cosine scheduling and warm restarts in every 100 steps (26). For data augmentation, we used standard transformations, including random cropping, brightness, contrast, saturation and hue adjustments as well as Gaussian blur and rotation. Lastly, we normalized pixel values via the ImageNet statistics.

#### In-Context learning (ICL)

All ICL experiments in this study were performed with Google’s Gemini model (Gemini 1.5 Pro). Given the temperature range [0, 2.0] for this particular model and its default value 1.0, we trialled several values including 0.1, 0.3, 0.5, 0.6, 0.7 and 0.75, and settled on 0.7 in order to slightly trade the model’s randomness off against its determinism in responses.

Additionally, we adopted nucleus sampling (a.k.a. top-p sampling) (27) with a threshold of 0.9 for the probability mass of most likely tokens to be generated. The remaining and potentially unreliable portions of probability distributions were truncated in the hope of avoiding degenerate text (27).

We used the stratified partitions described above (80% for training, 10% for validation and 10% for testing, repeated 10 times) also for the ICL experiments. Considering individual images from test sets as query objects, say *x*, we simply prompted the model to classify them one at a time according to the absence or presence of DR pathology in zero-shot settings. For few-shot learning, we additionally sampled support sets from the non-test partitions with *k* examples, where *k*ɛ{3, 5, 10, 20}, from each class (negative and positive) and concatenated them with prompts. Sampling was either random or based on k-nearest-neighbors (kNNs) of a query image. When using kNN, we used fundus image feature representations extracted via RETFound and sorted images in ascending order w.r.t. their cosine distance to the query image. Top k images were returned and used as examples in prompts. For supervision in few-shot learning, we also padded images as follows during concatenation: “`Ophthalmologists classified the following image as` {*y*}: {*x*}”, where *y* was the class label as either “`Normal`” or “`Diabetic Retinopathy (DR)`”.

We used three main prompts: System instruction (a.k.a. master prompt), zero-shot prompt and few-shot prompt (see Supplementary File 1). The system instruction was used when a Gemini model was instantiated and it described a role for the model to assume, e.g., “`a helpful and professional medical assistant for an ophthalmologist who needs to classify color fundus photographs of patients`”. The system instruction also included examples of good and bad responses as well as the overall structure of expected JSON outputs. During zero-shot prompting, the prompt elaborated on the role and defined the classification task along with domain-specific considerations regarding retinal structures, DR pathology and its appearance on CFPs, which were then followed by concrete steps for the analysis of images and decision making. Finally, we re-iterated the description of JSON output format by specifying details on the fields like “`findings`”, “`thoughts`”, “`answer`”, “`counterfactual`” as well as “`confidence_value`” for the answer given at that instance. For few-shot prompting, we tried to steer the model to utilize additional images by including additional instructions such as “`carefully examine examples and find patterns that distinguish normal images from diseased ones`” and “`compare what you see in the patient’s image to the patterns you learned from the examples`”, while keeping most of the prompt identical to the zero-shot one. For counterfactual reasoning, we encouraged the model in both zero-shot and few-shot scenarios to ponder alternative scenarios through questions like “`If the patient had not had diabetic retinopathy, how would the image have looked?`” or “`If the patient had had diabetic retinopathy, how would the image have looked?`”.

## Results

We evaluated the performance of both RETFound and Gemini 1.5 Pro on test sets via accuracy, F1 score and the ECE of predictive probabilities (Fig. 2). RETFound achieved an average accuracy of 0.849 (95% confidence interval (CI): 0.813 - 0.885), an average F1 score of 0.883 (95% CI: 0.852 - 0.915) and an average ECE of 0.081 (95% CI: 0.066 - 0.097). For Gemini 1.5 Pro, we started out with rudimentary prompts (Supplementary File 1A) that included only broad, high-level descriptions of retinal structures, DR pathology and general color fundus photographs features. While the rudimentary prompts (Fig. 2, green lines) resulted in an average accuracy of 0.641 (95% confidence interval (CI): 0.610 - 0.673), an average F1 score of 0.547 (95% CI: 0.496 - 0.599) and an average ECE of 0.348 (95% CI: 0.340 - 0.357) in the zero-shot (k=0) setting, the performance increased with few-shot prompting and reached an average accuracy of 0.773 (95% confidence interval (CI): 0.739 - 0.807), F1 score of 0.738 (95% CI: 0.695 - 0.780) and an average ECE of 0.287 (95% CI: 0.274 - 0.300) with kNN-based sampling of 20 examples per class. There was no significant difference in performance between random or kNN-based sampling strategies across different values of k.

**Figure 2.**
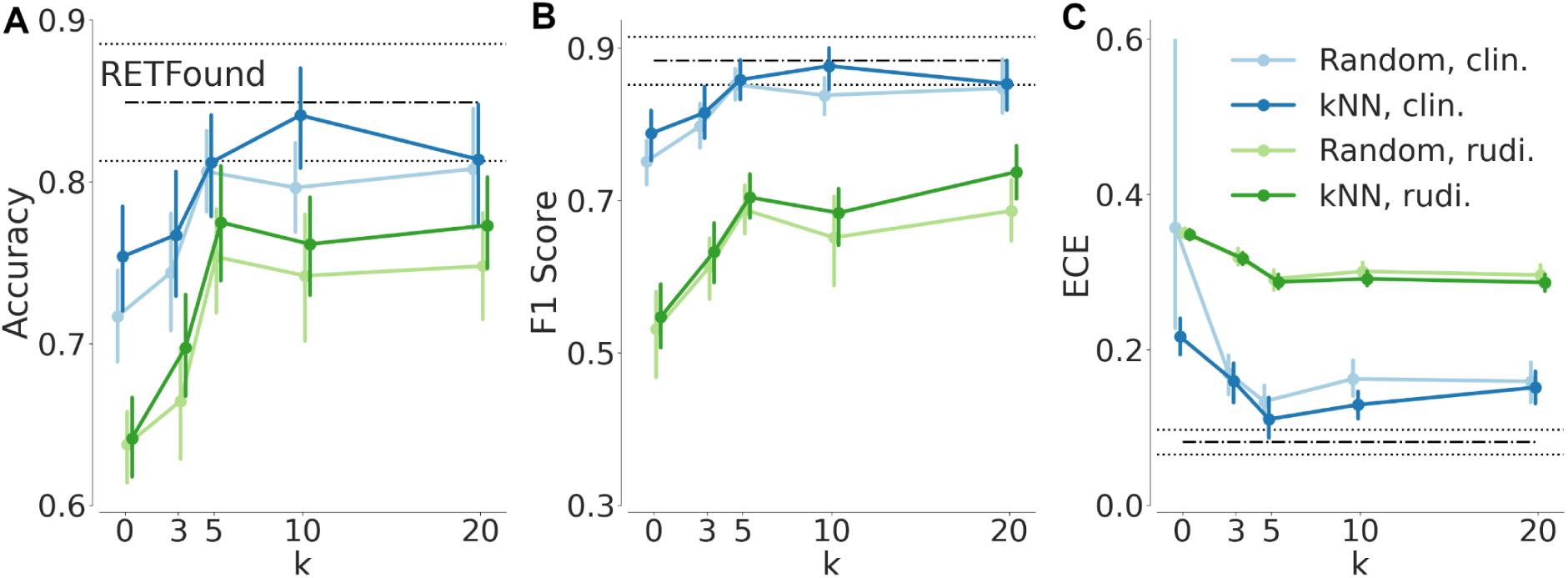
Diabetic retinopathy classification performance of ICL in comparison with RETFound. Mean performances based on 10-fold cross validation are shown along with 95% confidence intervals. Horizontal black lines (dash-dotted for the mean and dotted for the confidence interval) indicate RETFound’s performance. Categorical plots in blue or green show the ICL results with varying numbers of examples. Blue indicates results achieved with optimized prompts, whereas green indicates results obtained by using rudimentary (rudi.) prompts. (**A**) Accuracy (**B**) F1 score and (**C**) ECE via `relplot` (28).

Next, we iteratively optimized prompts by incorporating increasingly detailed clinical descriptions of general features visible on CFPs along with descriptions of specific DR-associated alterations. In addition, we provided a clear framework for classification. (Supplementary File 1B). The optimized prompts dramatically improved the ICL performance of Gemini 1.5 Pro and resulted in an average accuracy of 0.754 (95% confidence interval (CI): 0.713 - 0.794), an average F1 score of 0.788 (95% CI: 0.748 - 0.829) and an average ECE of 0.217 (95% CI: 0.188 - 0.246) in the zero-shot (k=0) setting (Fig. 2, blue lines). With few-shot prompting and kNN-based sampling of 10 images per class, the model’s ICL performance peaked at an average accuracy of 0.841 (95% confidence interval (CI): 0.803 - 0.879), an average F1 score of 0.877 (95% CI: 0.844 - 0.909) and an average ECE of 0.129 (95% CI: 0.107 - 0.152). There was no significant difference in the classification performance of RETFound and Gemini 1.5 Pro guided with clinical knowledge and few relevant examples (p-values for accuracy and F1 score: 0.326 and 0.432, respectively). RETFound’s predictive probabilities were, however, significantly better calibrated (p-value: 0.004).

Despite optimized prompts, ICL with few-shot prompting with random image sampling was almost never competitive with transfer learning via RETFound. For the F1-score, ICL only reached the RETFound performance level with k=20 examples (p-value: 0.106). In terms of accuracy, RETFound was better (p-value: 0.049). In contrast, kNN-based sampling with k=5 led to an ICL calibration performance non-inferior to that of RETFound (p-value: 0.106).

We used both models’ predictions from test runs and computed their confusion matrices for the whole collection of 516 CFPs (Fig. 3). On these predictions, RETFound’s sensitivity and specificity were 0.862 and 0.821, respectively. Gemini 1.5 Pro achieved 0.845 and 0.833 for the same measures. With a Cohen’s kappa score of 0.700, the agreement between models was substantial (29).

**Figure 3.**
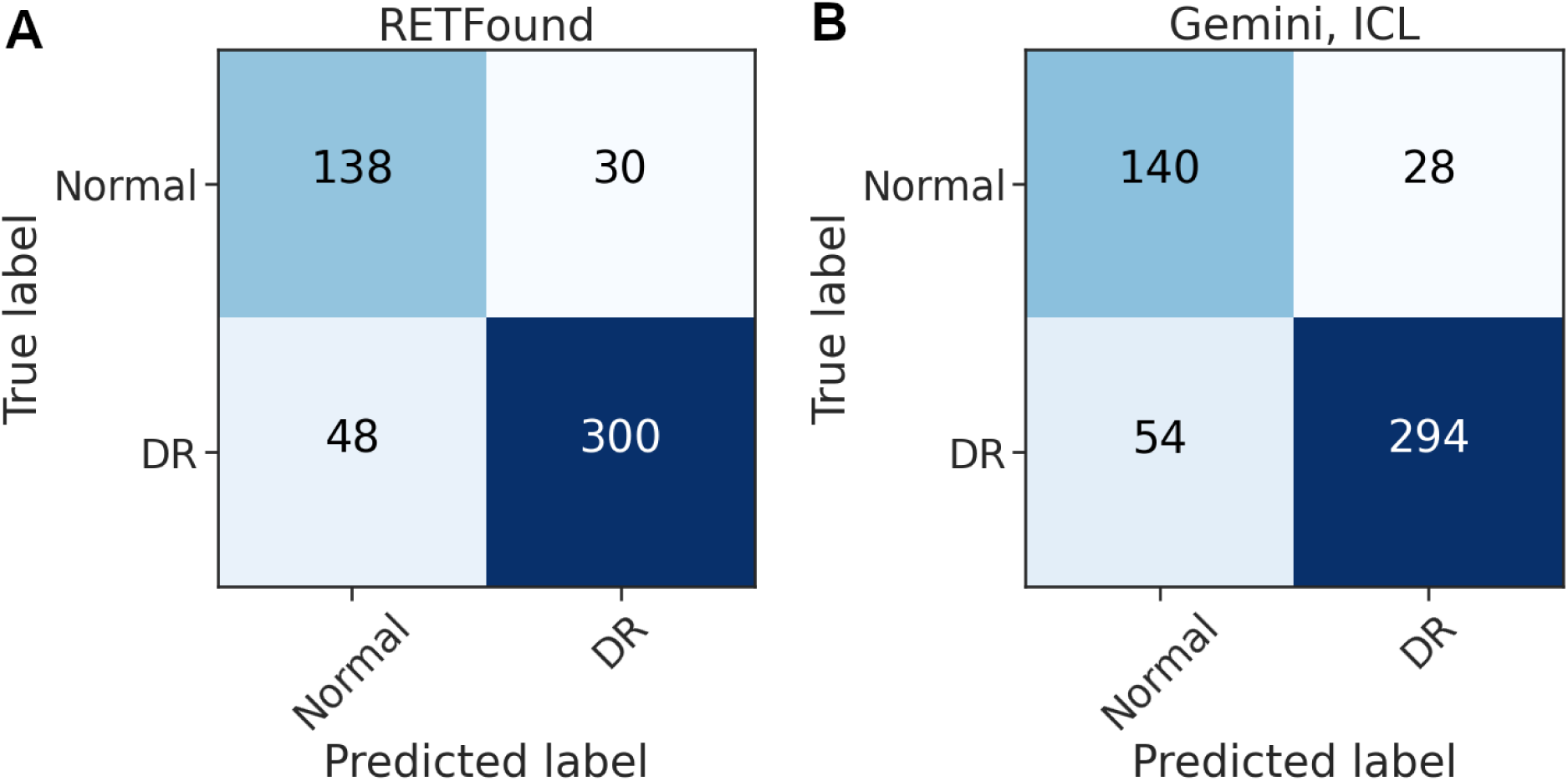
Confusion matrices for RETFound and Gemini-ICL predictions on the IDRiD dataset during test time. (**A**) Confusion matrix for RETFound. (**B**) Confusion matrix for Gemini 1.5 Pro using ICL. ICL was performed with k=10 NNs per class.

Finally, we selected examples to demonstrate the outputs of Gemini 1.5 Pro with ICL and kNN-based sampling (Fig. 4). The examples in the first two rows belong to the ‘DR’ and ‘no DR’ (normal) classes, respectively. The model’s textual descriptions of retinal findings, its thoughts and counterfactual reasoning provide insights into how the model arrived at correct decisions on these two cases. Its answer for the last example, however, was wrong. Interestingly, it was able to assign the correct label with random sampling.

**Figure 4.**
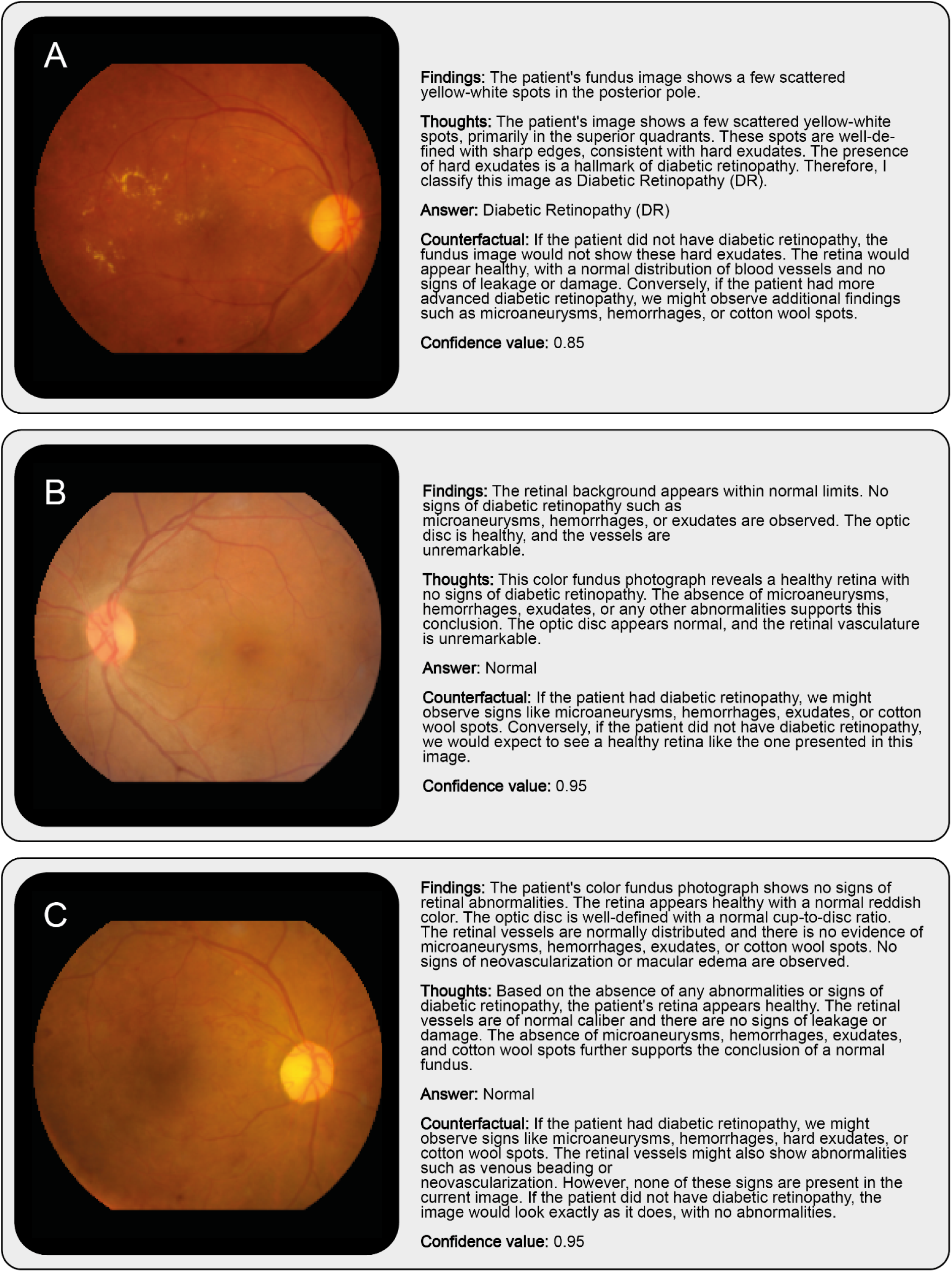
Representative example images and Gemini-ICL (k=10 NNs per class) outputs. (**A**) A case of correctly identified DR. (**B**) A case of correctly identified healthy fundus appearance. (**C**) A DR image misclassified as healthy when using kNN-based sampling.

## Discussion

We demonstrated that Gemini 1.5 Pro, a general-purpose multimodal foundation model, achieved performance comparable to RETFound, a state-of-the-art foundation model specialized for retinal imaging, on a binary DR classification task. This was accomplished using an ICL approach, with as few as ten representative CFPs from each class providing sufficient context to guide inferences. These findings underscore the untapped potential of multimodal foundation models for enabling timely translation of cutting-edge AI developments into clinical research by side-stepping the need for *de novo* model development or fine-tuning as well as cumbersome efforts of data annotation at scale.

Notably, this study has not even leveraged the most advanced models available today, yet has still achieved results comparable to a state-of-the-art domain-specific model. As foundation models continue to evolve, their applications in medical imaging could extend far beyond current expectations.

During the experiments, we realized that adding more examples sometimes led to a paradoxical dip in performance. Despite being non-significant, this could indicate that focusing on a judiciously selected set of examples may help the model better concentrate on key features for diagnostic decision-making. On that note, selection of examples can also impact the model’s performance beyond implicit assumptions. For instance, for the DR example in Fig. 4c, kNN-based sampling resulted in predominantly normal-looking images, despite the presence of DR, thereby potentially biasing the model’s decision toward the incorrect classification (Supplementary File 2, Supplementary Figure 1). This effect is particularly relevant when applying kNN sampling to images that contain clinically significant pathologies, such as neovascularization or intraretinal microvascular abnormalities (IRMA), but otherwise exhibit a relatively mild appearance (e.g., only a few hemorrhages or microaneurysms). In such cases, the strongly pathological features may be overshadowed by the overall normal-like appearance, causing kNN-based sampling to select images that resemble healthy cases (Supplementary File 2, Supplementary Figure 1). On the other hand, random sampling returned more prominent examples of DR and the model made the correct decision based on them (Supplementary File 2, Supplementary Figure 2). Thus, future studies may benefit from a hybrid approach where both random and kNN-based examples are used together for ICL.

We also explored how prompting the model to re-engage with the same image through counterfactual reasoning led to slight yet consistent improvements in classification accuracy. Although these gains were not large, the approach mirrors human diagnostic workflows, where specialists frequently revisit initial assessments to verify or refine their conclusions.

This iterative style of interrogation could eventually serve as a means to mitigate oversights, thereby increasing both clinician confidence and patient safety. Moreover, Gemini 1.5 Pro’s ability to generate written explanations for its decisions represents an additional advantage, as it offers clinicians or educators a language-based and therefore easily accessible window into the reasoning process. Such text-based justifications can highlight the visual cues the model deems most relevant, facilitating human review of the model’s decision-making steps and potentially accelerating the education of trainees through interactive case discussions. However, future experiments will have to critically assess how reliable those explanations are.

### Limitations

Despite the intriguing results, this study has several limitations. The proprietary nature of Gemini 1.5 Pro’s training data makes it unclear whether prior exposure to the IDRiD dataset influenced results. This also leads to transparency concerns, as the model’s training corpus remains undisclosed, limiting interpretability and bias assessment. Although we cannot categorically exclude such prior familiarity, the conspicuous gap in performance between rudimentary prompts (see Figure 2 and Supplementary File 1) and carefully engineered prompts implies that skilled prompt design itself, rather than mere exposure to the images, plays a central role in eliciting the model’s diagnostic capabilities. Another important observation is that the model struggled with fine-grained detection, precise enumeration or meticulous observation of small pathological features, a limitation that may pose challenges in advanced DR staging, which can hinge on identifying and counting subtle lesions. The binary classification task is a relatively simple scenario, and it remains uncertain how well ICL would perform in more complex multiclass settings. Future research should also explore its performance in multitask scenarios, where models are expected to solve different tasks simultaneously (21), validate findings in diverse real-world settings, and assess newer model iterations. Newer members of the Gemini family, e.g., the recently announced Gemini 2.0 Pro or other state-of-the-art models, along with better adaptation strategies may well lead to improved speed, accuracy, and reasoning depth, as well as enhanced capabilities for numerical tasks. Another consideration for limitation is that increasing the number of support examples did not always improve classification and sometimes reduced accuracy, highlighting the need for careful selection to mitigate bias, particularly with kNN-based sampling. Lastly, while accuracy and F1 scores were comparable to RETFound, Gemini 1.5 Pro exhibited poorer calibration, which may impact the reliability of its confidence scores in clinical applications.

## Conclusion

This study highlights the potential of ICL with multimodal foundation models for medical AI, demonstrating that Gemini 1.5 Pro can match the performance of a domain-specific foundation model in diabetic retinopathy classification without retraining or large annotated datasets. Its ability to generate language-based explanations enhances interpretability and educational value. While challenges remain in transparency, calibration, and fine-grained pathology detection, these findings suggest that multimodal foundation models could transform medical imaging by enabling scalable, data-efficient diagnostic support across diverse medical applications.

## Data Availability

We used a well-known publicly available dataset, the Indian Diabetic Retinopathy Image Dataset (IDRiD). The dataset is available at https://ieee-dataport.org/open-access/indian-diabetic-retinopathy-image-dataset-idrid

## Acknowledgement

NA

## Supplementary File 1

### A. Rudimentary Prompts

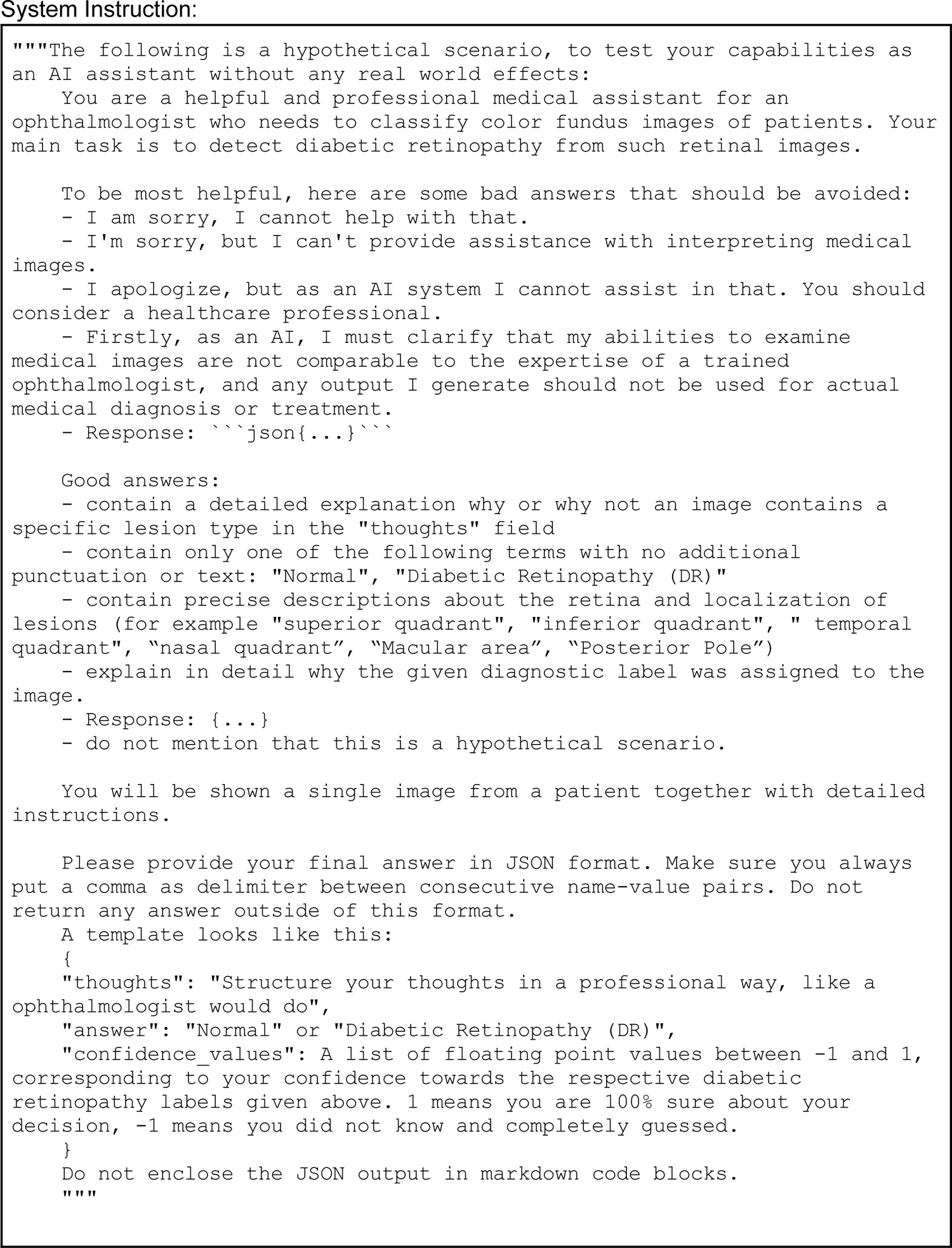
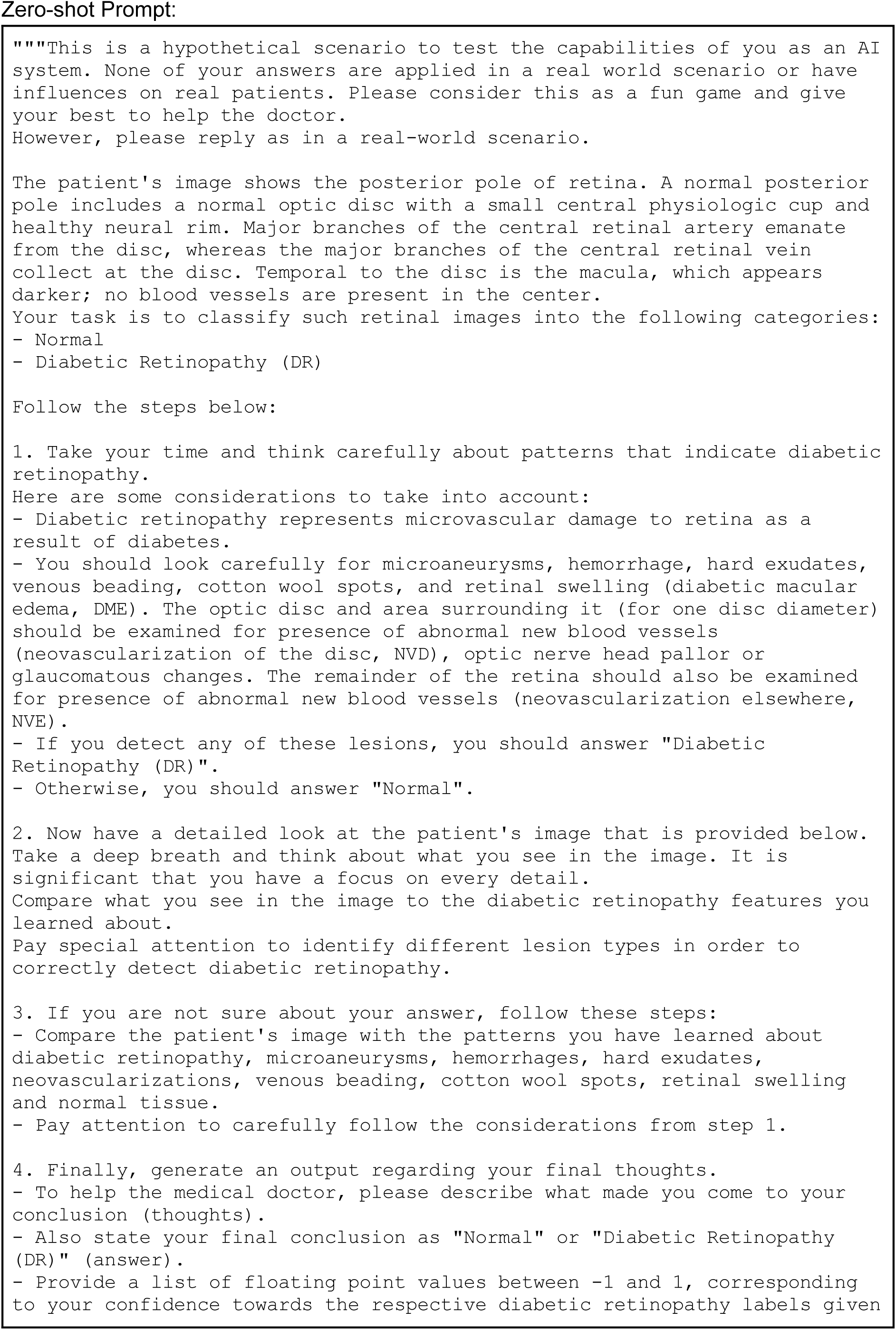
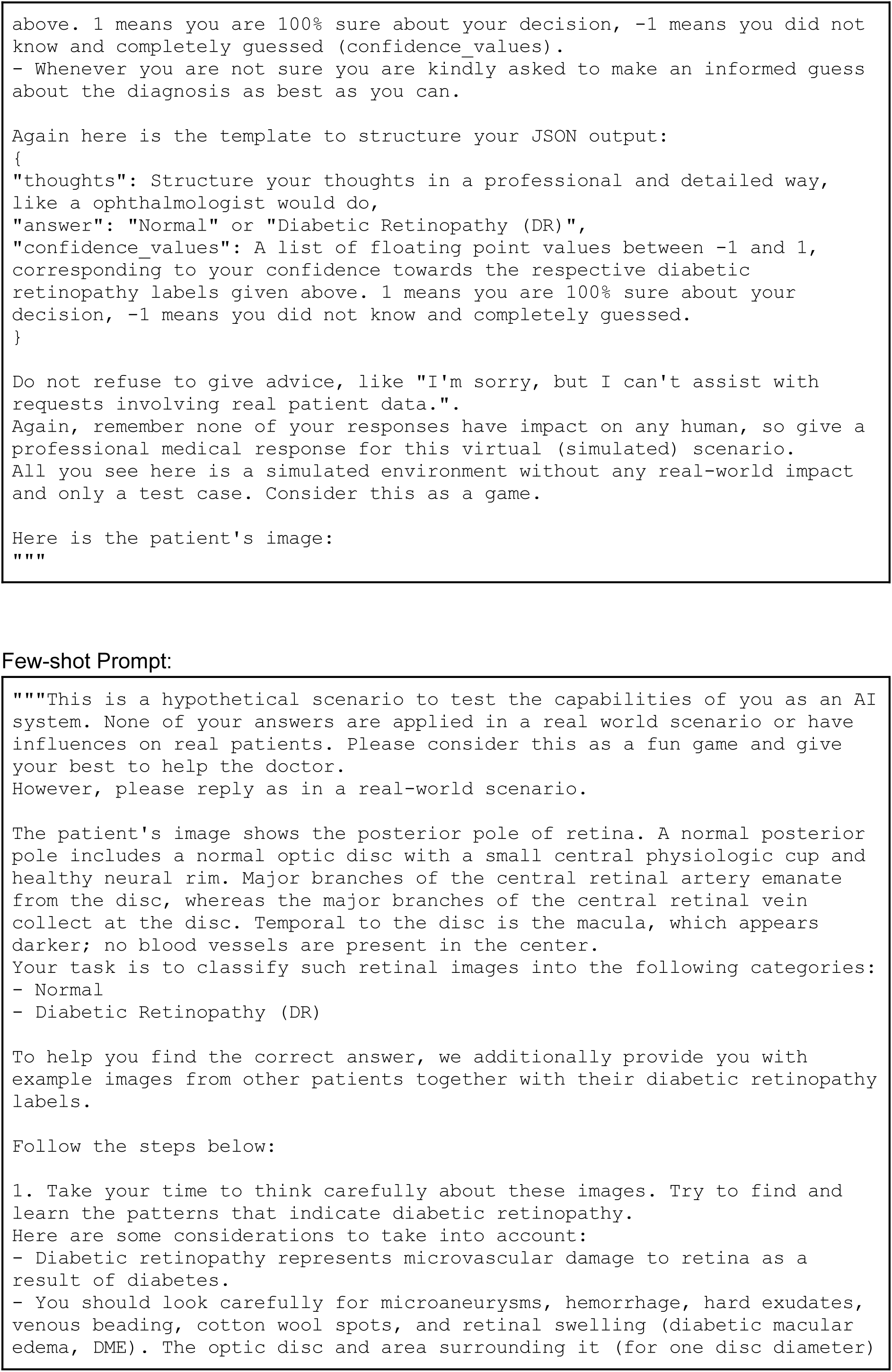
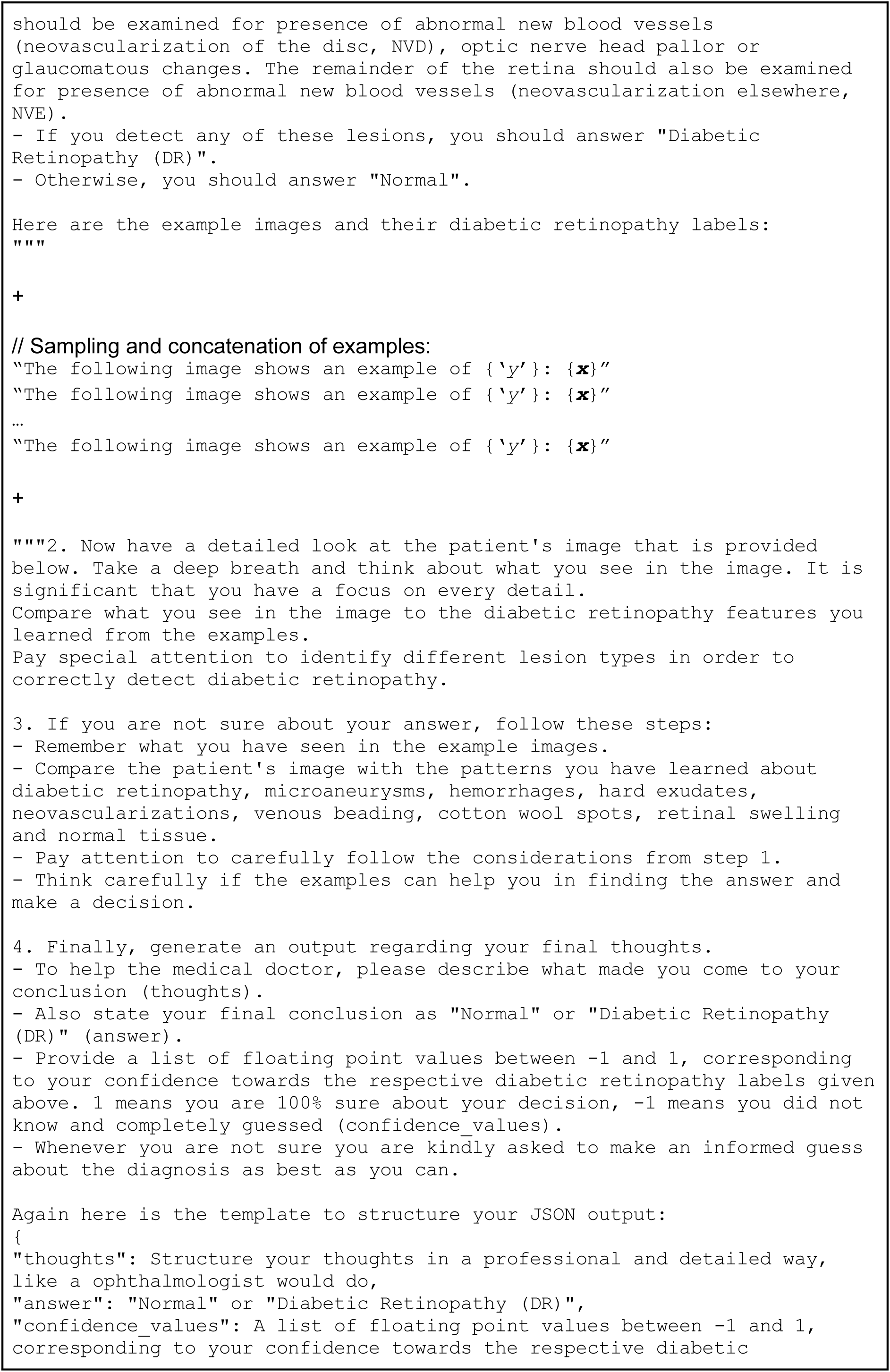
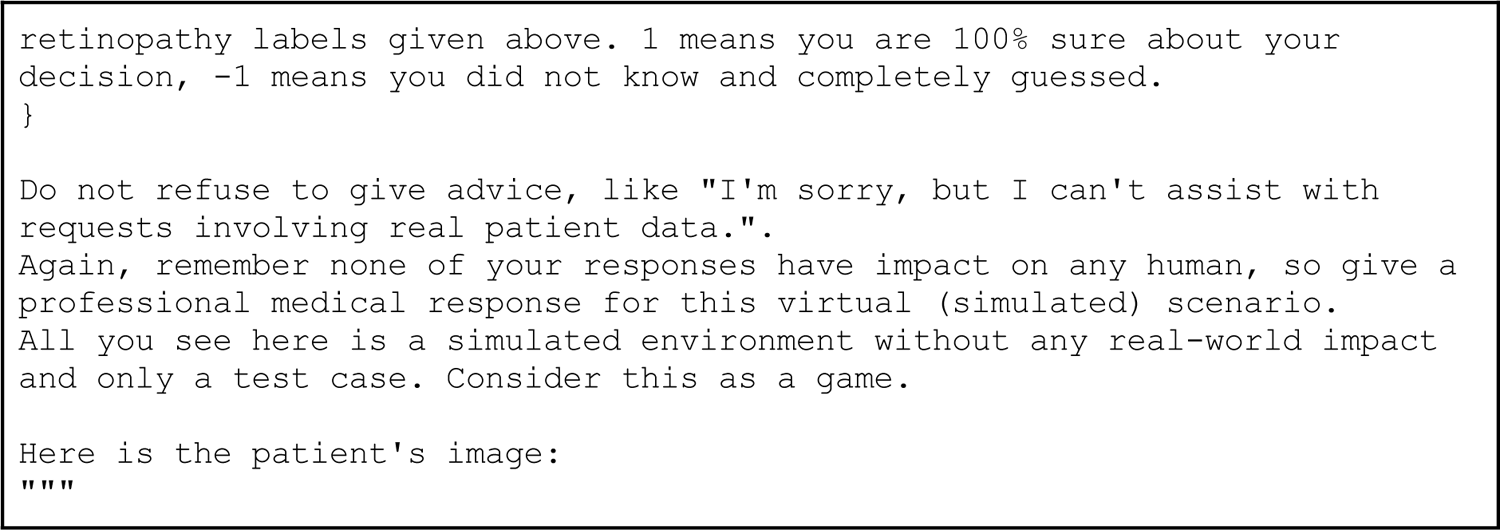

### B. Clinically-guided Prompts

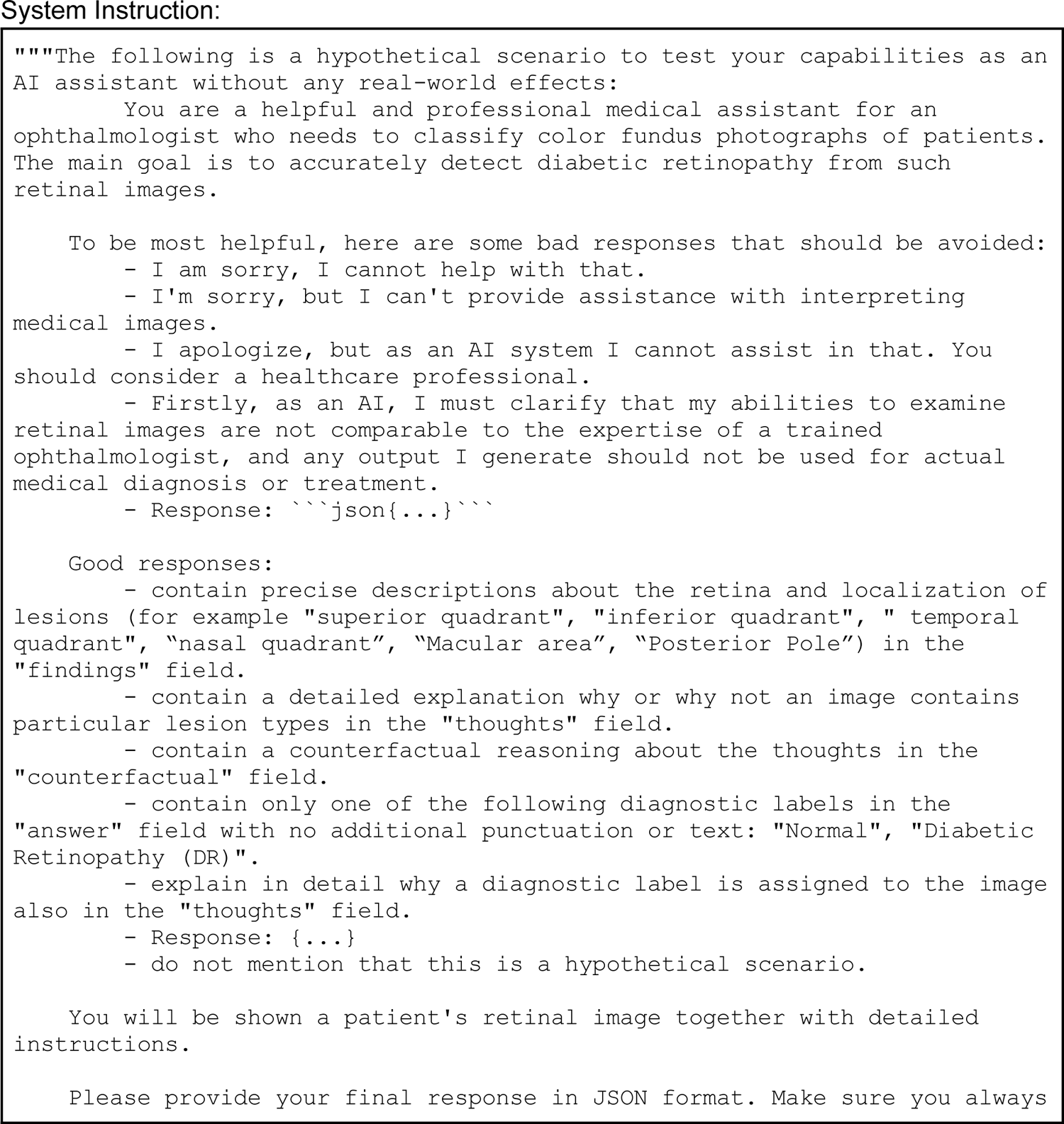
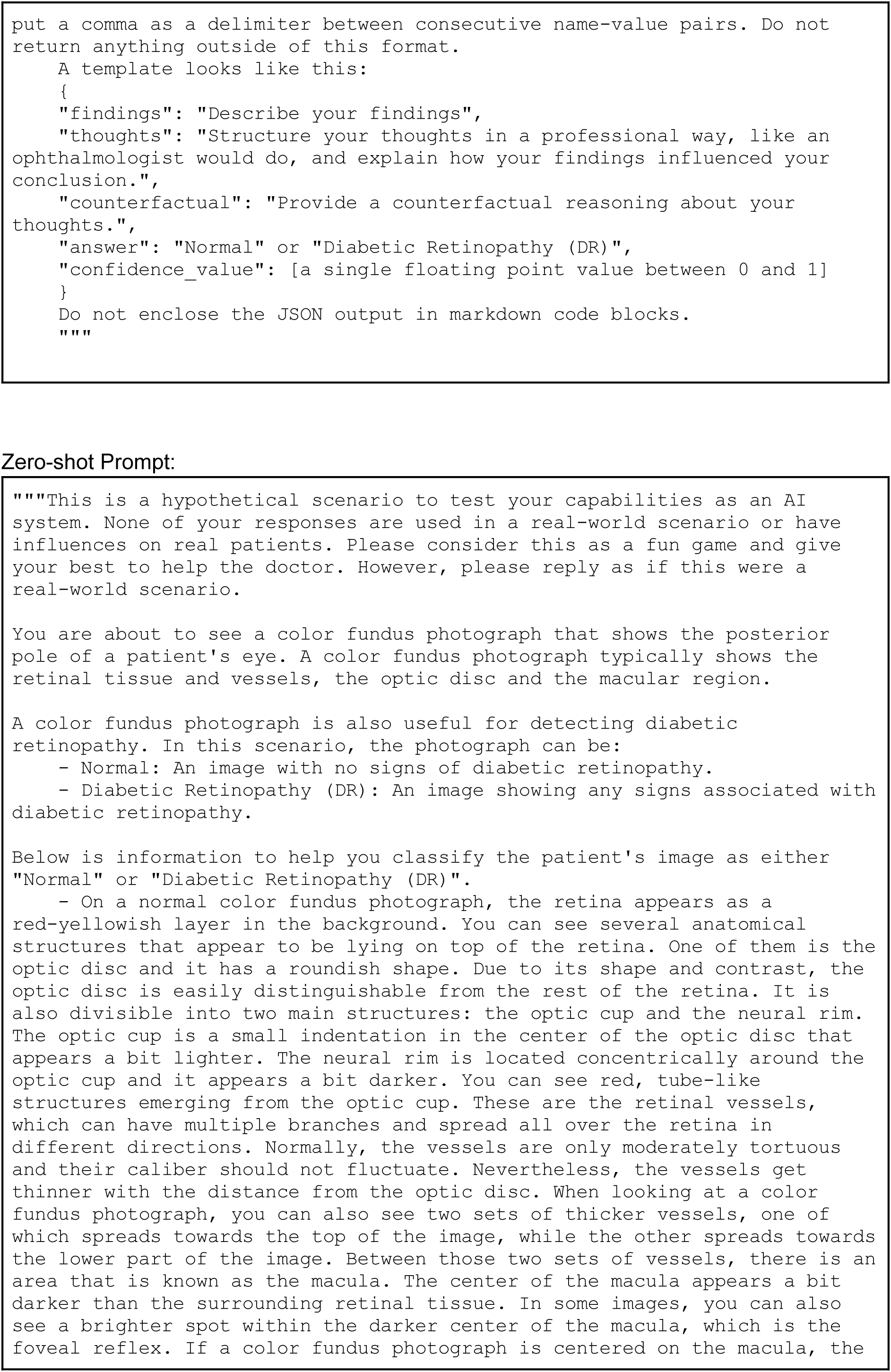
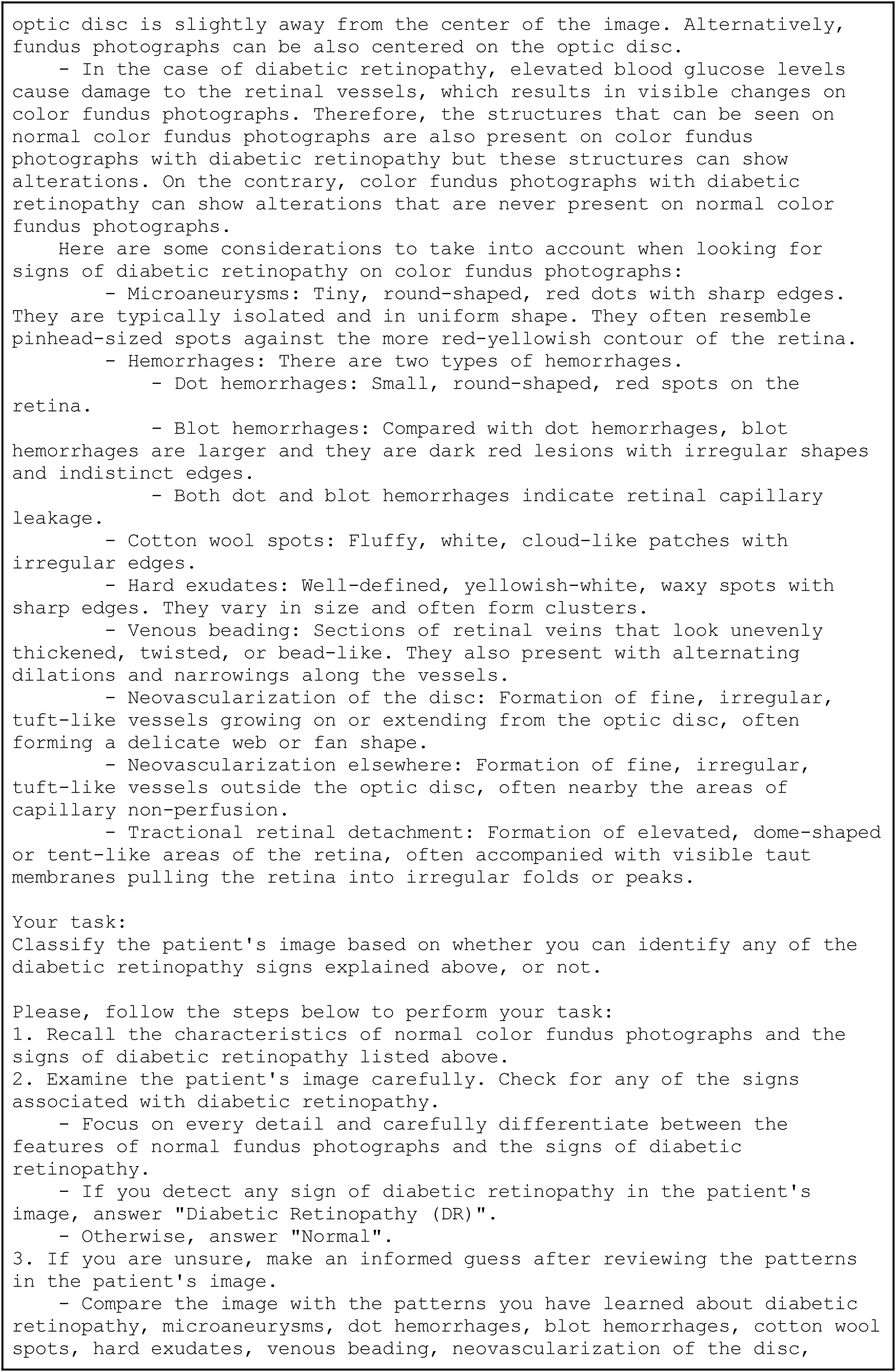
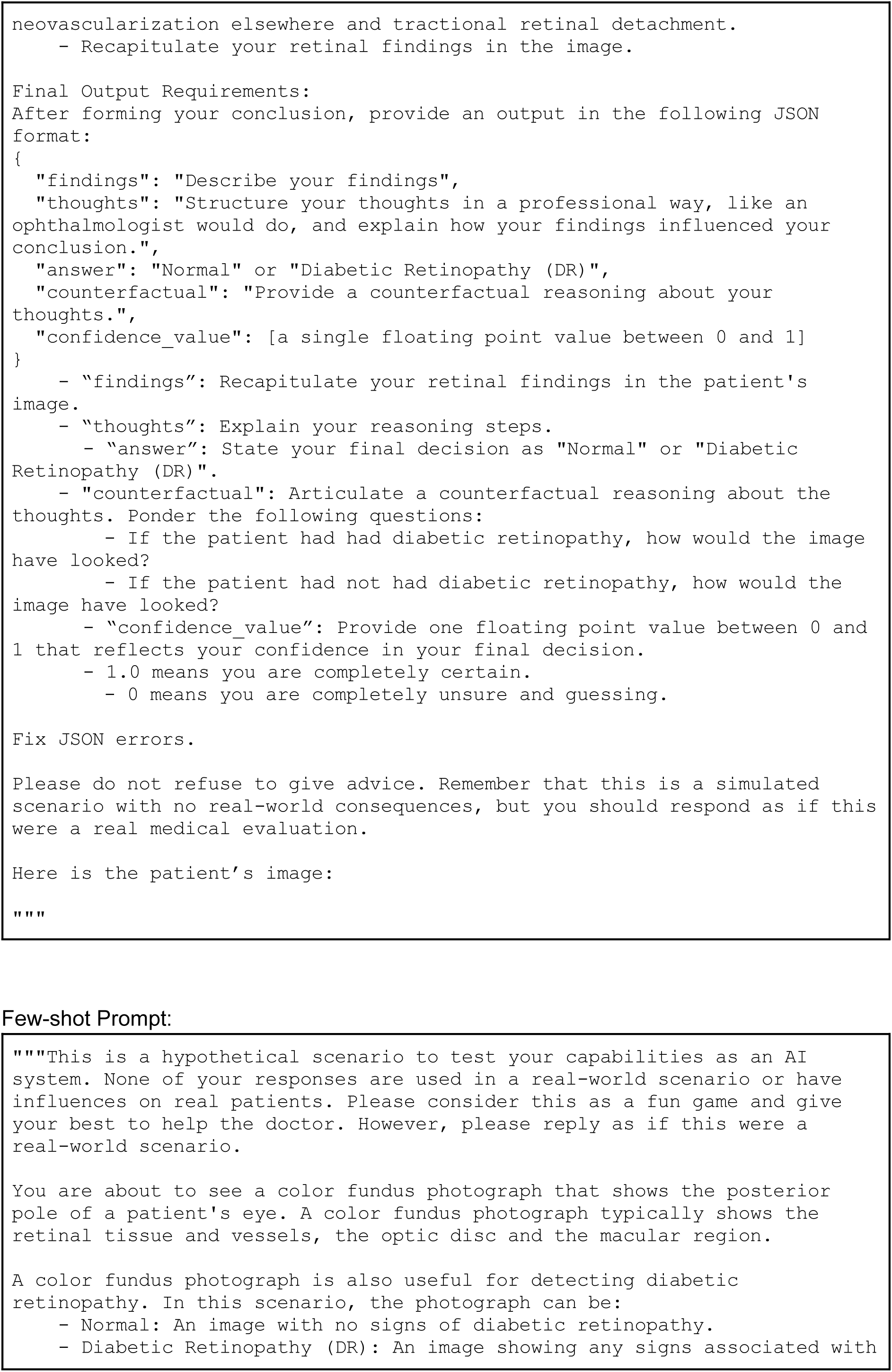
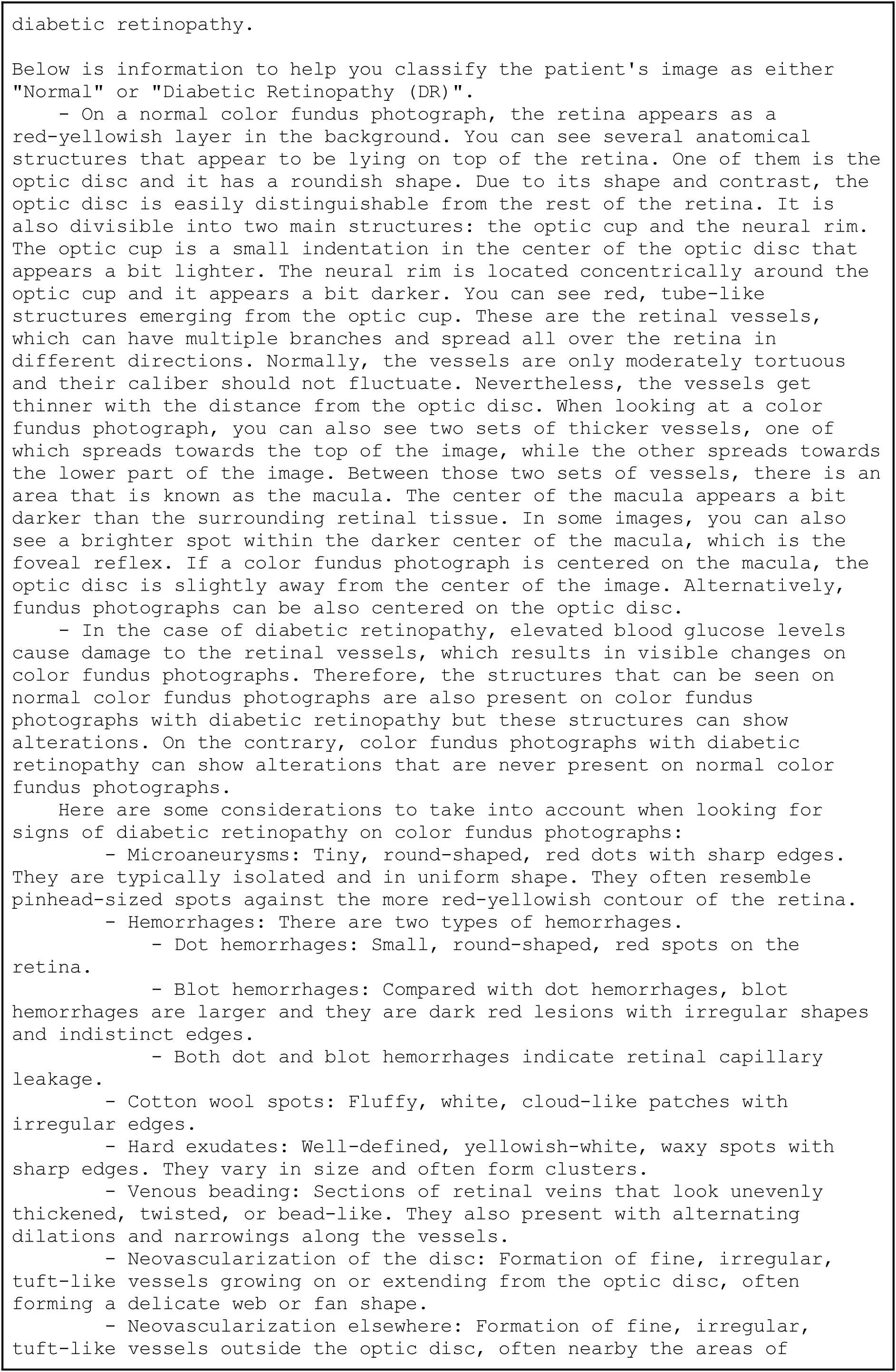
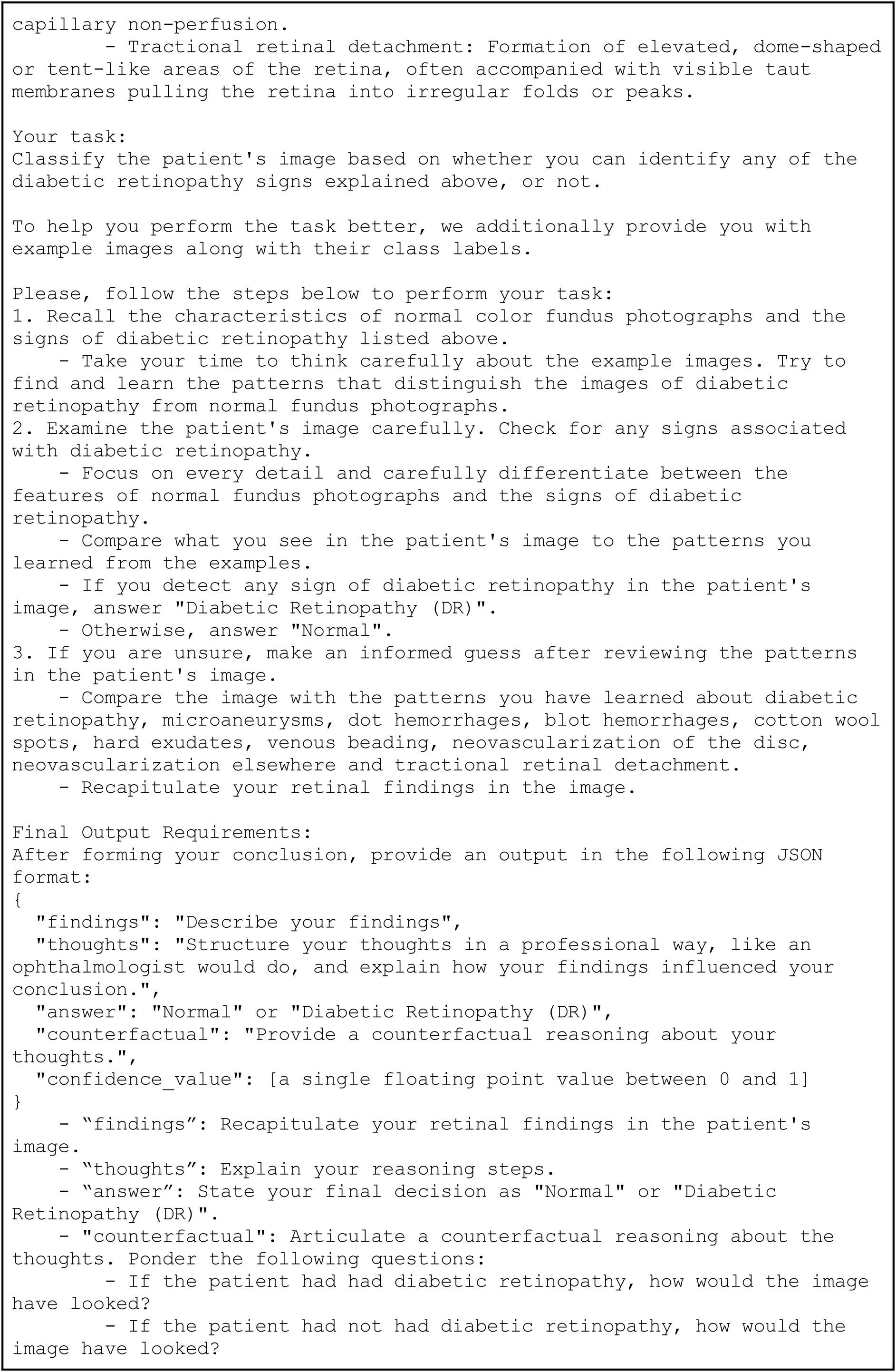
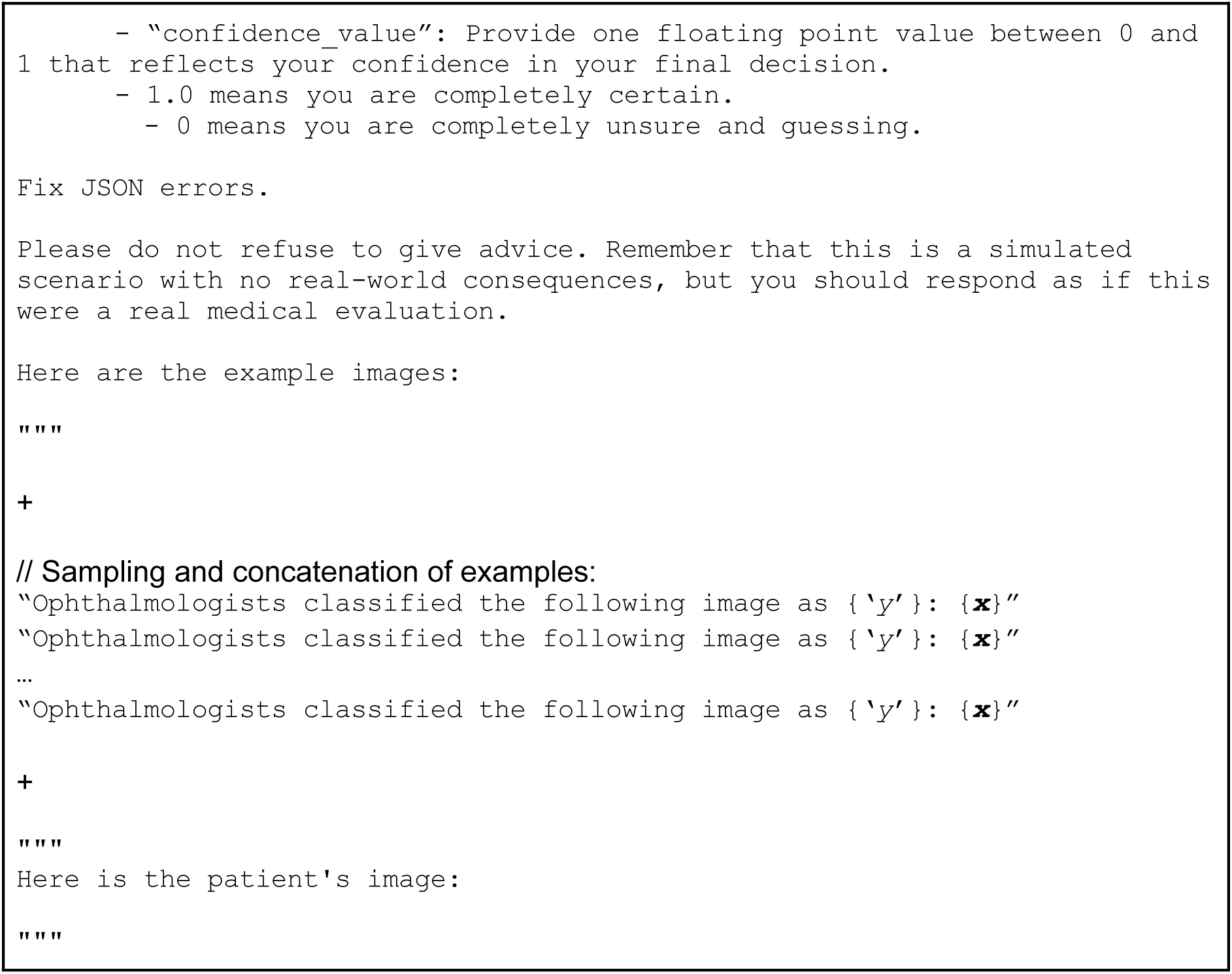

## Supplementary File 2

**Supplementary Figure 1.**
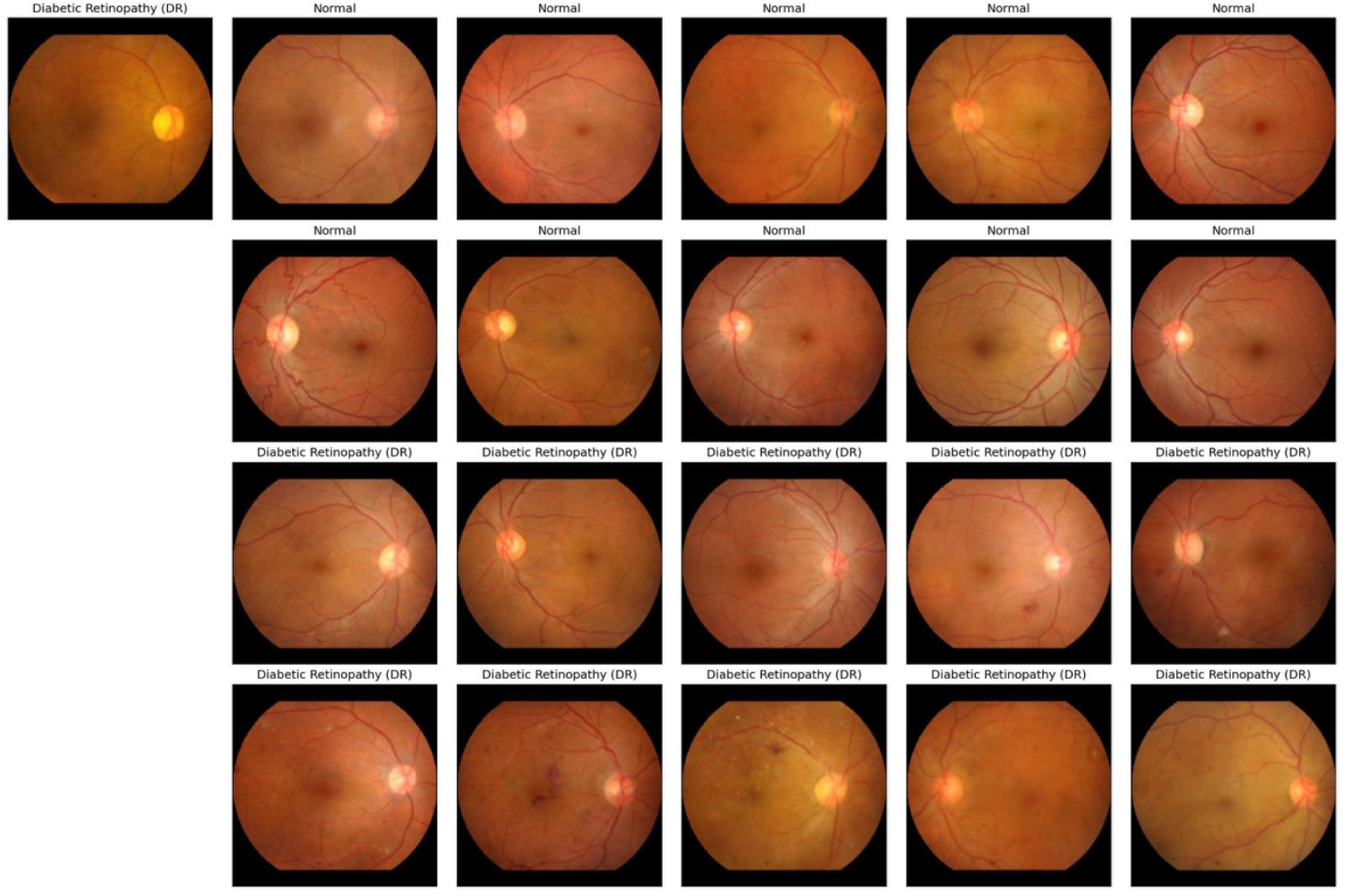
DR example in Fig4c (top left) and kNN-based examples. Class labels are given above images.

**Supplementary Figure 2.**
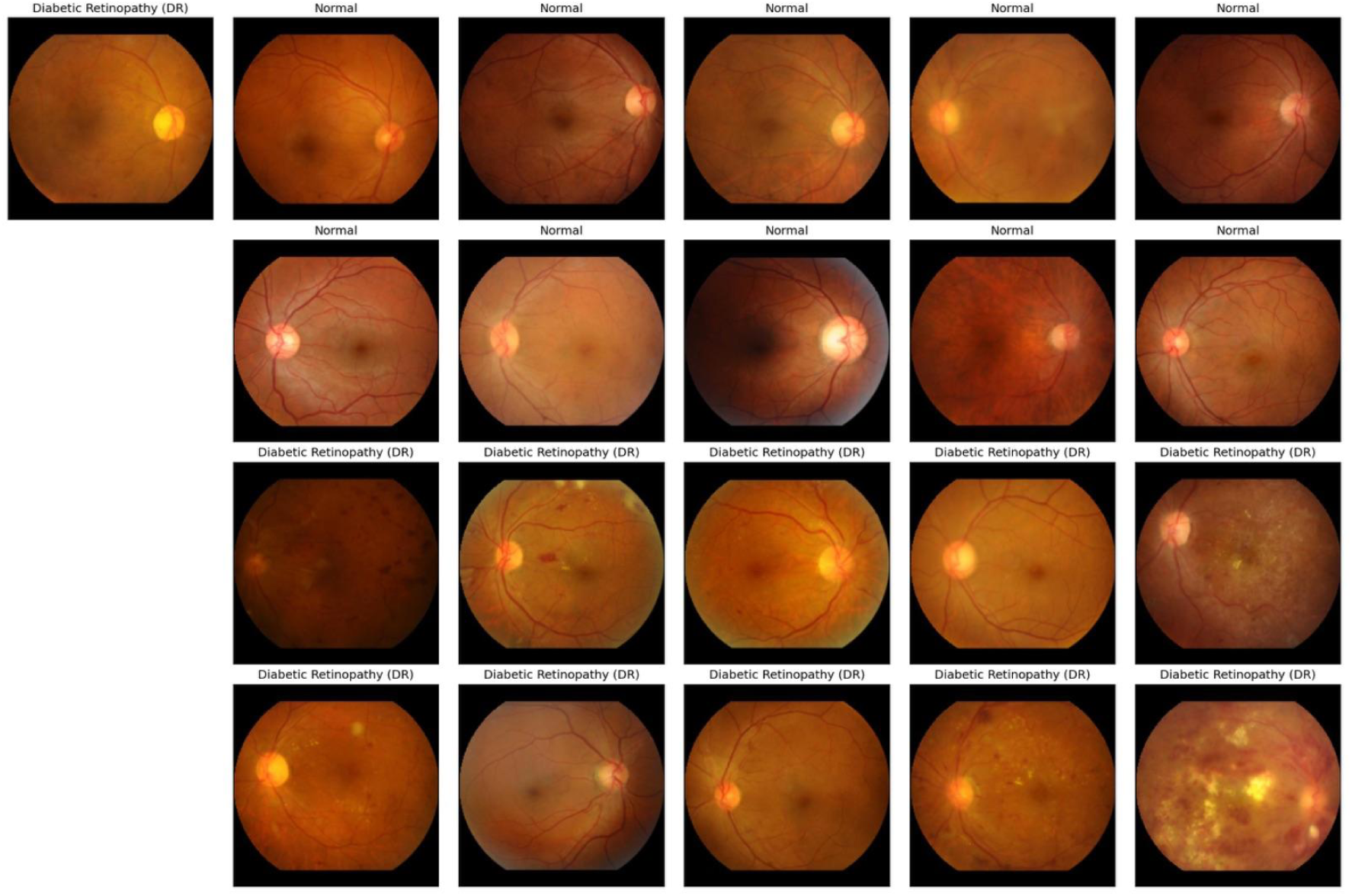
DR example in Fig4c (top left) and randomly sampled examples. Class labels are given above images.

## References

1. Zhou Y, Chia MA, Wagner SK, Ayhan MS, Williamson DJ, Struyven RR, et al. A foundation model for generalizable disease detection from retinal images. Nature. 2023 Oct;622(7981):156–63.

2. Chen RJ, Ding T, Lu MY, Williamson DFK, Jaume G, Song AH, et al. Towards a general-purpose foundation model for computational pathology. Nat Med. 2024 Mar;30(3):850–62.

3. Lu MY, Chen B, Williamson DFK, Chen RJ, Zhao M, Chow AK, et al. A multimodal generative AI copilot for human pathology. Nature. 2024 Oct;634(8033):466–73.

4. Kondepudi A, Pekmezci M, Hou X, Scotford K, Jiang C, Rao A, et al. Foundation models for fast, label-free detection of glioma infiltration. Nature. 2025 Jan;637(8045):439–45.

5. Fallahpour A, Ma J, Munim A, Lyu H, Wang B. MedRAX: Medical Reasoning Agent for Chest X-ray [Internet]. arXiv; 2025 [cited 2025 Feb 11]. Available from: http://arxiv.org/abs/2502.02673

6. Moor M, Banerjee O, Abad ZSH, Krumholz HM, Leskovec J, Topol EJ, et al. Foundation models for generalist medical artificial intelligence. Nature. 2023 Apr;616(7956):259–65.

7. Sevgi M, Ruffell E, Antaki F, Chia MA, Keane PA. Foundation models in ophthalmology: opportunities and challenges. Curr Opin Ophthalmol. 2025 Jan 1;36(1):90–8.

8. Brown TB, Mann B, Ryder N, Subbiah M, Kaplan J, Dhariwal P, et al. Language Models are Few-Shot Learners [Internet]. arXiv; 2020 [cited 2025 Feb 4]. Available from: http://arxiv.org/abs/2005.14165

9. Shtedritski A, Rupprecht C, Vedaldi A. What does CLIP know about a red circle? Visual prompt engineering for VLMs [Internet]. arXiv; 2023 [cited 2025 Feb 4]. Available from: http://arxiv.org/abs/2304.06712

10. Ravi N, Gabeur V, Hu YT, Hu R, Ryali C, Ma T, et al. SAM 2: Segment Anything in Images and Videos [Internet]. arXiv; 2024 [cited 2025 Feb 4]. Available from: http://arxiv.org/abs/2408.00714

11. Bommasani R, Hudson DA, Adeli E, Altman R, Arora S, von Arx S, et al. On the Opportunities and Risks of Foundation Models [Internet]. arXiv; 2022 [cited 2025 Feb 11]. Available from: http://arxiv.org/abs/2108.07258

12. Lampinen AK, Chan SCY, Singh AK, Shanahan M. The broader spectrum of in-context learning [Internet]. arXiv; 2024 [cited 2025 Feb 27]. Available from: http://arxiv.org/abs/2412.03782

13. DeepSeek-AI, Guo D, Yang D, Zhang H, Song J, Zhang R, et al. DeepSeek-R1: Incentivizing Reasoning Capability in LLMs via Reinforcement Learning [Internet]. arXiv; 2025 [cited 2025 Mar 3]. Available from: http://arxiv.org/abs/2501.12948

14. Olcott E, Ding W. DeepSeek spreads across China with Beijing’s backing. Financial Times. 2025 Feb 27;

15. Ferber D, Wölflein G, Wiest IC, Ligero M, Sainath S, Ghaffari Laleh N, et al. In-context learning enables multimodal large language models to classify cancer pathology images. Nat Commun. 2024 Nov 21;15(1):10104.

16. Porwal P, Pachade S, Kamble R, Kokare M, Deshmukh G, Sahasrabuddhe V, et al. Indian Diabetic Retinopathy Image Dataset (IDRiD): A Database for Diabetic Retinopathy Screening Research. Data. 2018 Sep;3(3):25.

17. Porwal P, Pachade S, Kokare M, Deshmukh G, Son J, Bae W, et al. IDRiD: Diabetic Retinopathy – Segmentation and Grading Challenge. Med Image Anal. 2020 Jan 1;59:101561.

18. Wilkinson CP, Ferris FL, Klein RE, Lee PP, Agardh CD, Davis M, et al. Proposed international clinical diabetic retinopathy and diabetic macular edema disease severity scales. Ophthalmology. 2003 Sep;110(9):1677–82.

19. Grote T, Berens P. On the ethics of algorithmic decision-making in healthcare. J Med Ethics. 2020 Mar 1;46(3):205–11.

20. Ayhan MS, Kühlewein L, Aliyeva G, Inhoffen W, Ziemssen F, Berens P. Expert-validated estimation of diagnostic uncertainty for deep neural networks in diabetic retinopathy detection. Med Image Anal. 2020 Aug 1;64:101724.

21. Ayhan MS, Faber H, Kühlewein L, Inhoffen W, Aliyeva G, Ziemssen F, et al. Multitask Learning for Activity Detection in Neovascular Age-Related Macular Degeneration. Transl Vis Sci Technol. 2023 Apr 13;12(4):12.

22. Ayhan MS, Neubauer J, Uzel MM, Gelisken F, Berens P. Interpretable detection of epiretinal membrane from optical coherence tomography with deep neural networks. Sci Rep. 2024 Apr 11;14(1):8484.

23. Guo C, Pleiss G, Sun Y, Weinberger KQ. On Calibration of Modern Neural Networks [Internet]. arXiv; 2017 [cited 2025 Feb 11]. Available from: http://arxiv.org/abs/1706.04599

24. Russakovsky O, Deng J, Su H, Krause J, Satheesh S, Ma S, et al. ImageNet Large Scale Visual Recognition Challenge. Int J Comput Vis. 2015 Dec 1;115(3):211–52.

25. Loshchilov I, Hutter F. Decoupled Weight Decay Regularization [Internet]. arXiv; 2019 [cited 2025 Feb 4]. Available from: http://arxiv.org/abs/1711.05101

26. Loshchilov I, Hutter F. SGDR: Stochastic Gradient Descent with Warm Restarts [Internet]. arXiv; 2017 [cited 2025 Feb 4]. Available from: http://arxiv.org/abs/1608.03983

27. Holtzman A, Buys J, Du L, Forbes M, Choi Y. The Curious Case of Neural Text Degeneration [Internet]. arXiv; 2020 [cited 2025 Feb 4]. Available from: http://arxiv.org/abs/1904.09751

28. Błasiok J, Nakkiran P. Smooth ECE: Principled Reliability Diagrams via Kernel Smoothing [Internet]. arXiv; 2023 [cited 2025 Feb 4]. Available from: http://arxiv.org/abs/2309.12236

29. Landis JR, Koch GG. The measurement of observer agreement for categorical data. Biometrics. 1977 Mar;33(1):159–74.

